# Assessing the reliability and validity of pictorial-assisted 24-hour recall for measuring hand hygiene and child faeces disposal: a cross-sectional study in Malawi

**DOI:** 10.1101/2024.08.11.24311806

**Authors:** Olivier Rizk, Sarah Bick, Blessings White, Kondwani Chidziwisano, Robert Dreibelbis

## Abstract

Whilst improving hygiene and sanitation behaviours is key to cost-effective and sustainable WASH interventions, measuring behaviour change remains a challenge. This study assessed the validity and reliability of pictorial 24-hour recall (P24hR), a novel method using unprompted recall of past activities through pictures, compared to structured observation for measuring handwashing with soap (HWWS) and safe child faeces disposal in rural Malawi. Data were collected from 88 individuals across 74 households in Chiradzulu district using both methods over a two-day period, with the recall period of the P24hR corresponding to the period of structured observation completed the previous day. Results showed poor agreement between P24hR and observations in detection of hygiene opportunities and behaviours. P24hR under-reported handwashing opportunities when frequency was high and over-reported them when frequency was low. The 95% limits of agreement for handwashing opportunities estimated through Bland-Altman analysis (-7.62 to 4.89) were unacceptably wide given median 5 opportunities observed per participant. P24hR also over-reported HWWS and safe child faeces disposal, and kappa statistics indicated agreement no better than by chance. Structured observation remains the preferred method for measuring hygiene behaviours despite its known limitations, including potential reactivity bias.

## Introduction

Interventions to improve hand hygiene in domestic settings are associated with a 30% reduction in diarrheal diseases among children under the age of five (Wolf et al., 2022) and a 17% reduction in acute respiratory infections (Ross et al., 2023). Our estimates of the potential health benefits of hygiene interventions, however, are associated with exposure to – rather than adoption of – hand hygiene interventions. This is because measuring hygiene behaviour remains a challenge (Egreteau, 2017; Schmidt et al., 2019), with few validated and reliable methods for measuring behaviours available. Contaminated hands are a critical pathway for exposure to a range of environmentally transmitted pathogens (Wagner et al., 1958) and quantifying and measuring hand hygiene behaviour is a key part of exposure risk assessment (Kwong et al., 2020), intervention design and evaluation (Amon-Tanoh et al., 2021), and understanding individual and population-level health risks (Wolf et al., 2019).

Among the methods used to measure behaviour, structured observation is often considered the gold standard due to its ability to measure behaviours as they occur (Biran et al., 2008; Schmidt et al., 2019). However, it is resource-intensive and can be seen as intrusive or inappropriate for certain behaviours. Most importantly, direct observation can result in reactivity from participants, in which case the validity of estimates is limited (Ram et al., 2010).

Proxy measures – or indirect measures of behaviour – can also be used in hygiene and sanitation research. They are often operationalized as the presence of necessary materials or infrastructure to enable a specific behaviour (Schmidt et al., 2019). For example, the presence of a handwashing facility is used as a proxy measure for hand hygiene behaviour (Biran et al., 2008; Joint Monitoring Program, 2022) based on global estimates suggesting individuals are almost two times as likely to wash hands with soap after faecal contact events when both soap and water are available. Proxy measures are convenient as data can be rapidly collected and at a low cost, but their accuracy may be limited (Biran et al., 2008; Briceño et al., 2014).

Self-reporting tools are commonly used to measure behaviour. These tools are inexpensive, quick and require little expertise to put in place or use (Schmidt et al., 2019). However, self-reported hygiene and sanitation behaviours are often unreliable due to biases, including recall bias. Several studies have shown poor agreement between reported and observed hygiene behaviour (Chidziwisano et al., 2020; Curtis et al., 1993; Manun’Ebo et al., 1997; Stanton et al., 1987).

Pictorial 24h recall (P24hR) has been suggested as a novel methods to measure hygiene and sanitation behaviours (Schmidt et al., 2019). P24hR measures behaviours through facilitated recall of past activities with pictures and a diary sheet. P24hR is a validated method to measure dietary intake, with photos and pictures assumed to increase the accuracy of reporting compared to unfacilitated recall (Lazarte et al., 2012; National Cancer Institute, 2023). P24hR has been used to evaluate various handwashing interventions (Tidwell et al., 2019; Tidwell et al., 2020). In a study in India (Schmidt et al., 2019), researchers found that P24hR of handwashing behaviour was more closely aligned with direct observation data than reported hand hygiene. However, comparisons were made between two different study groups rather than compared among the same individuals; more information on the validity and reliability of P24hR is needed to further assess its utility in measuring hygiene behaviours.

The aim of this study was to assess the agreement of P24hR compared to direct observation for pre-selected hygiene and sanitation behaviours and determine the validity and reliability of P24hR. By measuring the same set of behaviours with different methods, we provided useful information regarding the measurement properties of P24hR compared to structured observation.

## Materials and methods

This field-based cross-sectional study compared the agreement between measured prevalence of hand washing with soap (HWWS) and safe child faeces disposal practices in a sample of rural households in Chiradzulu district, Malawi. Target behaviours were measured using both structured observations and P24hR in the same participants over a two-day period, with the recall period of the P24hR corresponding to the period of direct observation completed the previous day.

### Study setting and sampling

This study was conducted as part of a larger research and learning collaboration between the London School of Hygiene and Tropical Medicine and the Malawi University of Business and Applied Sciences. Chiradzulu is situated in the southern region of Malawi and is sub-divided into 8 Traditional Authorities (TA) (Figure 1). Villages included in this study were selected from a roster of villages present in TA-Likoswe and TA-Mpama. Both TAs were part of a community-based sanitation promotion programme implemented by the NGOs World Vision and Water For People the year before data collection.

**Figure 1.**
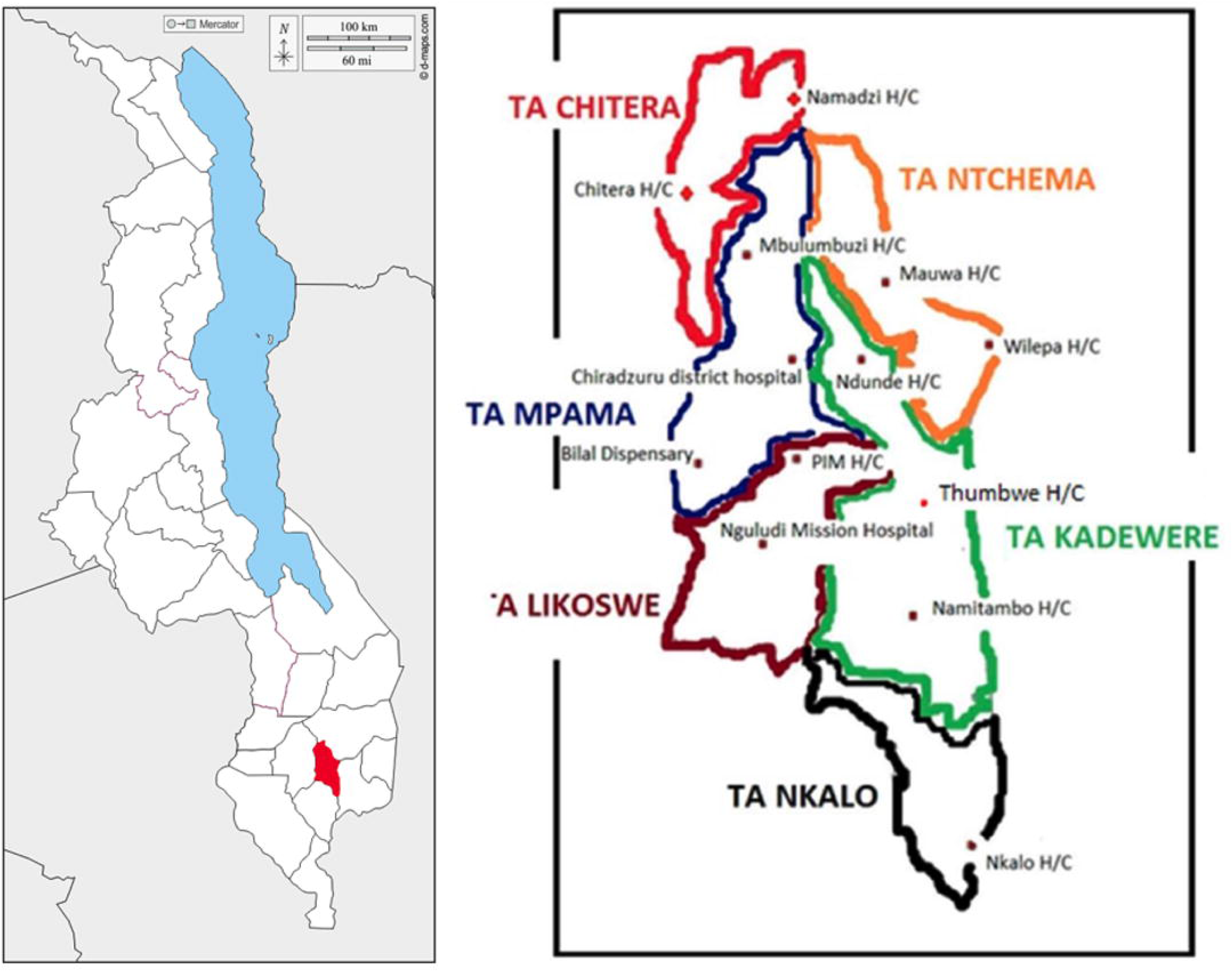
Maps of Malawi and Chiradzulu district. Left: Map of Malawi with Chiradzulu district highlighted in red (Adapted from (D-Maps.com, 2023)). Right: Map of Chiradzulu district and its traditional authorities taken from (Kazembe, 2018).

We aimed to enrol approximately 75 households based on the range typically used for agreement studies (Han et al., 2022). Sampling was completed in 13 villages across the study area, six from TA-Likoswe and seven from TA-Mpama, with each village contributing six households to the final sample size. Within villages, we approached every sixth household from a pre-defined starting point. The household inclusion criterion was the presence of a child under five years of age at the time of the observations, to ensure the possibility of observing child faeces disposal practices.

Within each household, we recruited up to 3 individuals to participate in study activities. Participants were adult (over 18 years old) residents of the household. In instances where households contained more than three adult residents present, the adults contributing most to childcare and household activities were selected in priority.

### Tool development and implementation

Detailed tool development is described in Appendix A. In brief, a list of daily activities was developed and adapted to local context, resulting in a list of 41 discrete activities. Each activity was translated into a pictorial image reflecting that specific activity (Appendix B). Daily routines were organized around 5 temporal periods – early morning (waking until breakfast), morning (breakfast through lunch), afternoon (after lunch until sunset), evening (sunset until the evening meal) and night (evening meal until bedtime). Pilot testing found that participants were able to map their reported activities to using the activity cards to the organized daily diary.

### Data collection

Data collection consisted of 8 staff who had prior experience with direct observation and water, sanitation and hygiene research. Field teams were organized into two teams: six observers and two enumerators conducting P24hR (henceforth ‘interviewers’). Observers were different from interviewers to reduce the risk of bias. The six observers were female, as it was easier for them to be allowed in homes where females were mostly present. After obtaining approval by village chiefs and collecting appropriate consent from participating household members, each household was visited twice over a two-day consecutive period to conduct direct observations (day 1) and P24hR (day 2).

Observations lasted six hours and began in the morning (around 7:30 am), when most household activities took place in the study population. Participants were all observed at the same time. Observers would generally sit in the yard, where many activities take place. If a participant left the household, observers would remain with the other participants. Opportunities for handwashing and their associated behaviour were recorded for all participants. Handwashing opportunities were pre-defined as: after going to the toilet, after taking children for defecation, after cleaning children after defecation, after disposing of children’s faeces, before washing food, before preparing food, before serving food, after tending to animals (Appendix A). For each opportunity, observers could record one of the following hand hygiene activities: no handwashing; handwashing with ash, mud or soil; handwashing with water only or handwashing with soap. Observers also recorded any child defecation event and the faeces disposal method: in the latrines, buried, in the open or in the garbage. Each of the six observers visited one household per day for a total of six households observed per day.

The administration of P24hR was completed the next day and took on average 20-30 minutes per participant. Participants were introduced to the 41 pictures and the diary sheet, explained how to use them to describe their activities in the past 24 hours, and then given time to complete the diary sheet independently. Interviewers would help if participants had difficulty identifying pictures or time periods. After the diary sheet was completed, interviewers would go through the participants’ day, one activity at a time, asking if they had forgotten anything. Finally, interviewers would manually record each activity and take a picture of the completed diary sheet. Each of the two interviewers visited three households per day for a total of six households per day.

The data from direct observations, pictorial 24h recall and household surveys were recorded on Android tablets with forms produced using the online platform KOBO Toolbox. Data were encrypted and uploaded daily to a secured server.

### Statistical analysis

All statistical analyses were conducted in Stata version 18 (StataCorp, College Station, TX, USA). The data collected during observations and P24hR were matched for each participant using a unique identifying code and the corresponding 6-hour observation period was isolated within the 24 hours of pictorial recall data for comparison (early morning and morning). Observed and reported opportunities and behaviours were extracted for each participant, for both HWWS and safe child faeces disposal.

The number of handwashing opportunities was a count variable totalling all opportunities for handwashing defined above. Handwashing opportunities that occurred in rapid succession in either the observation or P24hR data were treated as a single hand hygiene opportunity, for example ‘Washing food’ immediately followed by ‘Preparing food”. The number of HWWS events associated with an opportunity was originally constructed as a count variable, however, given the low rates of HWWS in both methods, we constructed a binary variable of any recorded HWWS associated with a handwashing opportunity during the period of interest.

Due to the low number of child defecation events, the count of events was converted into a binary variable representing any child defecation event during the period of interest. The binary variable of safe child faeces disposal was also defined as any safe disposal practices following child defecation as defined by the World Health Organization (buried or disposed in latrines) reported or observed during the period of interest (World Health Organization and United Nations Children’s Fund, 2006).

Inter-method agreement for the count outcome (number of handwashing opportunities, modelled as a continuous variable) was evaluated using the Bland-Altman method (Bland and Altman, 1986). Bland-Altman analyses plot the differences in values obtained by two methods against the respective mean values. The mean difference between the two methods, referred to as the bias, indicates the extent to which the methods diverge. The standard deviation of the bias is used to estimate limits of agreement (LOA), which act as a reference interval between which 95% of the data should lie. An advantage of Bland-Altman plots is that they allow to simultaneously assess reliability and validity of the methods relatively to each other (Montenij et al., 2016) and standard approaches are recommended for when data violate distributional assumptions (Bland and Altman, 1999).

Inter-method agreement for binary variables was evaluated using kappa statistics (Cohen, 1960). This method measures agreement between two methods compared to expected agreement by chance alone. Kappa statistics below 0 indicate agreement worse than by chance; values equal to 0 indicate agreement no better than chance, and values between 0 and 1 reflective of increasing agreement (Landis and Koch, 1977). Additionally, results were analysed using McNemar’s test to assess the symmetry in performances between the two methods based on marginal totals, providing an estimate or over- or under-reporting (Curtis et al., 1993; Manun’Ebo et al., 1997; McNemar, 1947).

Using direct observation at the reference group, we also compared the sensitivity and specificity of P24hR methods. True positives were defined as target behaviours reported by both P24hR and direct observations; target behaviours reported by P24hR but not observed were considered false positives. True negatives were defined as the absence of target behaviour in both P24hR and observation data; target behaviours not reported by P24hR but capturing during observations were classified as false negative. Sensitivity, specificity, and positive and negative predictive values were calculated to compare the two methods (Guitart et al., 2021; Trevethan, 2017).

### Ethical considerations

This study was approved by National Committee on Research in the Social sciences and Humanities in Malawi (Protocol No. P.01/23/718) as well as the Ethical Review Committee at the London School of Hygiene and Tropical Medicine (LSHTM MSc Ethics Ref: 28743). Informed consent was obtained in all households before beginning direct observations and confirmed either through their signature or a thumbprint if the participant was illiterate. In the case of an illiterate participant, the presence of a literate individual co-signing as an independent witness was also required.

## Results

### Sample Characteristics

In total, 88 individuals across 74 households participated in both structured observations and pictorial 24h recalls. Selected characteristics are presented in Table 1. In some of the smaller villages (<35 households), finding six households with a child under five was not always possible. Due to the time limitations, seven households without children under five were included to meet sample size requirements.

**Table 1.** Characteristics of participants and households.

|  |  |
| --- | --- |
| **Characteristics of participants** | n (%) or mean (SD) |
| n | 88 |
| Female | 80 (91%) |
| Age | 35.0 (14.5) |
| Education |  |
| Primary | 56 (63.6%) |
| Secondary | 29 (33.3%) |
| **Characteristics of households** |  |
| n | 74 |
| Household residents, median (IQR) | 5 (4, 6) |
| Presence of child < 5 years | 67 (90.5%) |
| Age of child < 5 years, mean (SD) | 2.7 (1.2) |
| Electricity | 11 (14.9%) |
| Mobile phone | 55 (74.3%) |
| Monthly income (MWK) |  |
| <K10,000.00 | 9 (12.2%) |
| K10,000.00 to K20,000.00 | 19 (25.7%) |
| K20,000.00 to K30,000.00 | 15 (20.3%) |
| K30,000.00 to K40,000.00 | 15 (20.3%) |
| K40,000.00 to K50,000.00 | 5 (6.7%) |
| >K50,000.00 | 11 (14.8%) |
| Water source |  |
| Unprotected well | 1 (1.3%) |
| Borehole or tubewell | 71 (96.0%) |
| Piped into compound, yard, plot | 2 (2.7%) |
| Sanitation facility |  |
| No toilet or neighbour's toilet (not shown) | 8 (10.8%) |
| Flush / pour flush | 1 (1.3%) |
| Pit latrine with slab | 32 (43.2%) |
| Pit latrine without slab | 33 (44.6%) |
| Handwashing facility |  |
| Mobile object reported (bucket / jug / kettle / tippy tap) | 11 (14.9%) |
| No handwashing place in dwelling/yard/plot | 63 (85.1%) |
| Presence of soap in the household | 46 (62.2%) |

### Measurement of handwashing opportunities and behaviours

P24hR detected 412 (median 4 per participant) total handwashing opportunities compared to 531 in structured observations (median 5 per participant) (Table 2). Differences between the two methods in counts of total opportunities per participant were consistent with a normal distribution (Figure 2; p = 0.20).

**Figure 2.**
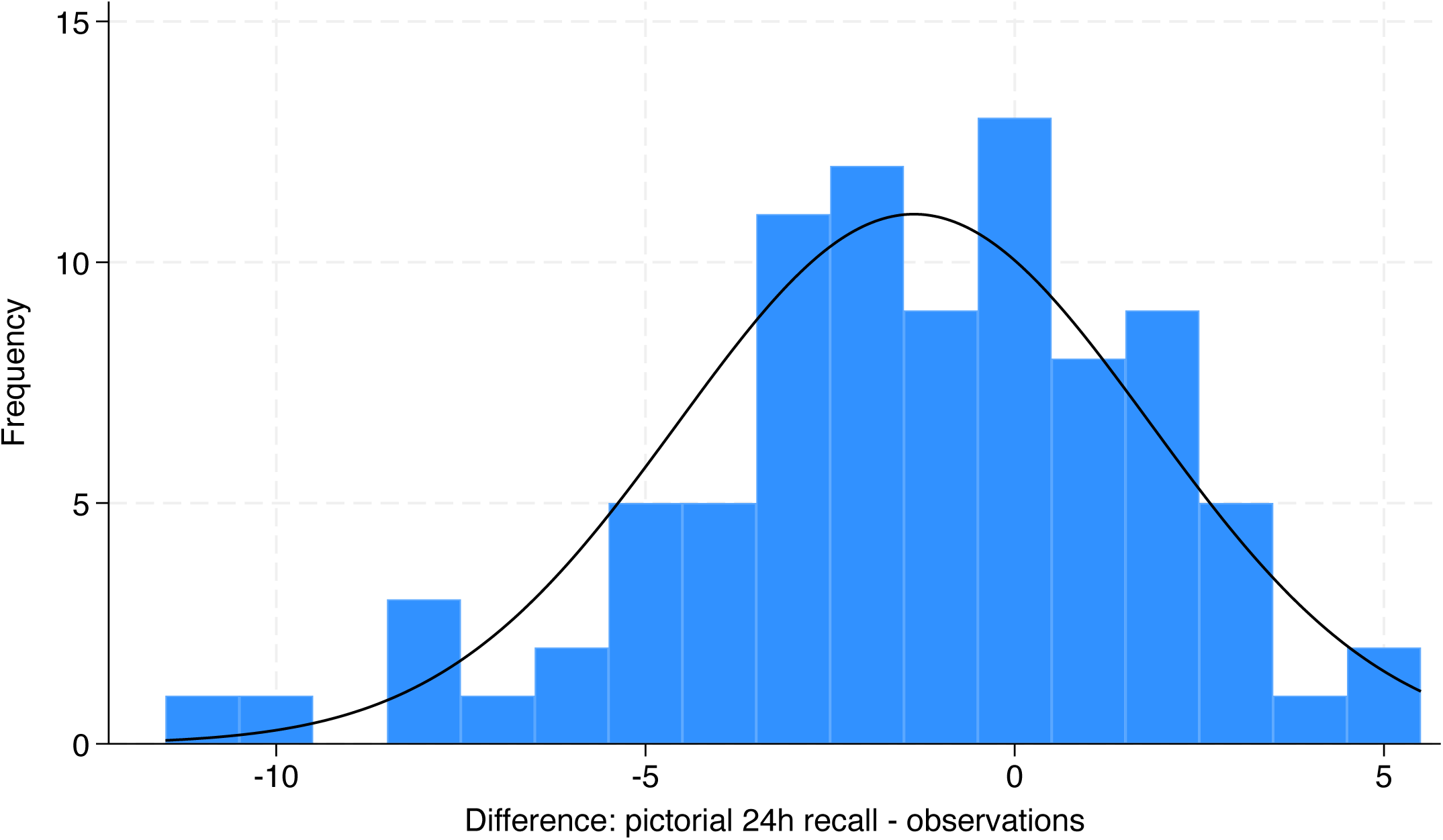
Histogram: difference in total number of handwashing opportunities measured by pictorial 24h recall and structured observation with normal density function overlaid.

**Table 2.** Observed and reported hygiene and sanitation opportunities and practices.

| Measurement method | Opportunities for HWWS | Opportunities per participant, median (IQR) | HWWS practiced (%) | Opportunities for child faeces disposal | Safe child faeces disposal (%) |
| --- | --- | --- | --- | --- | --- |
| Structured observations | 531 | 5 (3.5, 8.5) | 7 (1.3) | 6 | 5 (83.3) |
| P24hR | 412 | 4 (3, 6) | 29 (7.0) | 16 | 15 (93.8) |

Using a classical BA approach, bias was -1.36 (95% confidence interval (95%CI): -2.04, - 0.69), indicating under-reporting by P24hR, while the limits of agreement (LOA) extended from -7.62 to 4.89. Testing for the required assumptions for a classical analysis revealed that 7/88 observations (8.0%) laid beyond LOA and proportional bias was present as shown in Figure 3. Consequently, LOA were calculated using standard regression methods (Figure 3). For low average values, P24hR over-reported opportunities for handwashing, while for high average values, the method under-reported opportunities. P24hR was more precise when the average of opportunities was low compared to high averages as indicated by narrower LOA and data points closer to the line of equality.

**Figure 3.**
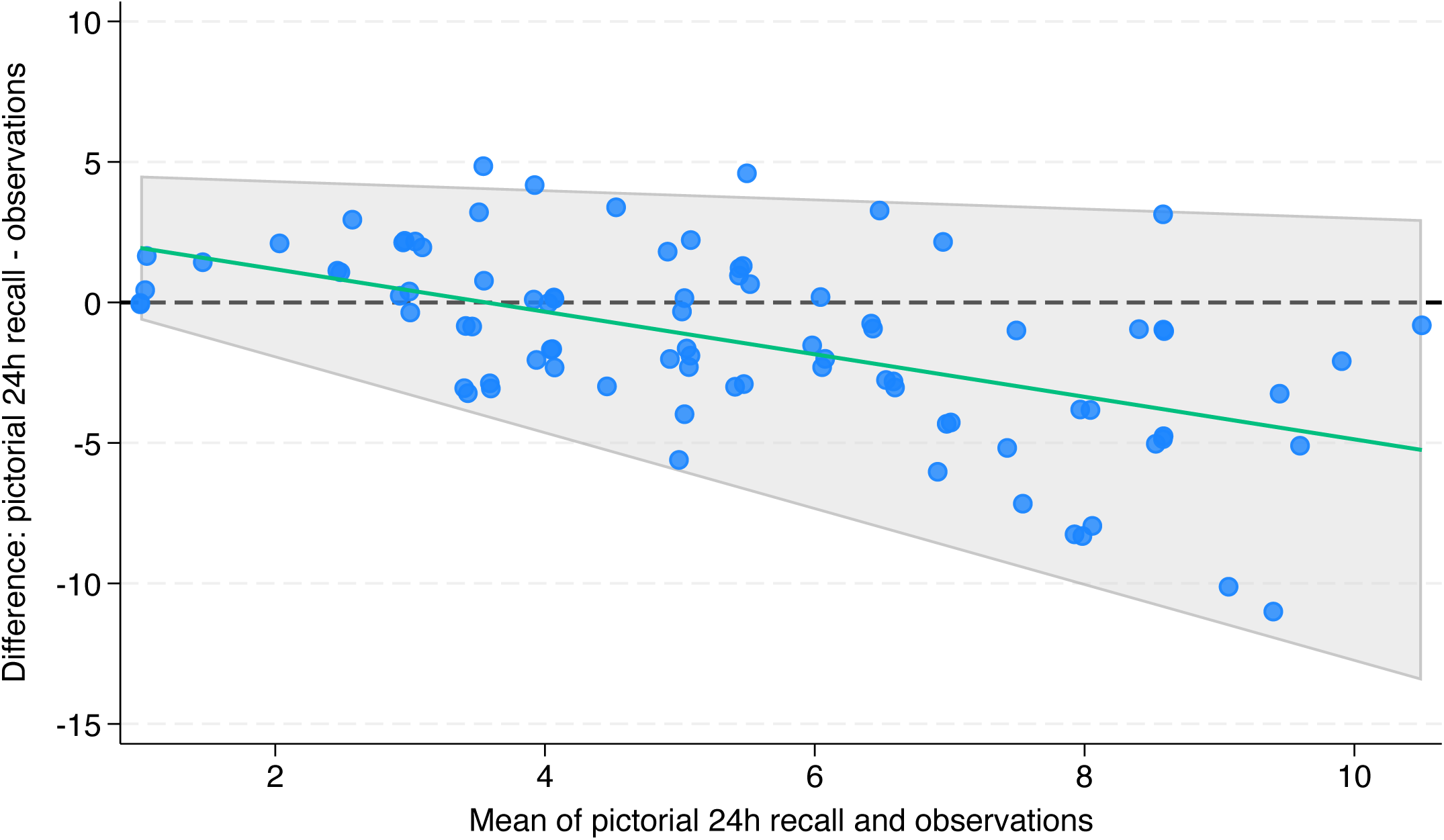
Bland-Altman plot of the number of handwashing opportunities with regression-based bias and limits of agreement. Bland-Altman plot of difference in number of opportunities for handwashing measured by P24hR and observations against the mean number of opportunities recorded by the two methods. Bias represented by a solid green line. Limits of agreement (mean difference +/- 2 SD) are shown by the shaded grey section. Bias is estimated by y = 2.70 - 0.757 * ((observations + P24hR)/2. Lower LOA is estimated by y= -0.719 - 1.35 * ((observations + P24hR)/2). Upper LOA is estimated by y = 4.67 - 0.164 * ((observations + P24hR)/2). Overlapping points separated by jitter effect.

Handwashing with soap was observed at 7 of the 531 opportunities (1.3%) while participants reported HWWS at 29/412 (7%) of opportunities identified in the P24hR (Table 2). Kappa statistic of presence of any HWWS was close to zero, indicating agreement no better than by chance (Table 3). Due to low rates of observed behaviour, a binary variable of any reported or observed HWWS was created for each participant. McNemar’s test of this binary variable gave strong evidence that the marginal prevalence of HWWS at any key moment differed between the two methods.

**Table 3.** Evaluation of validity of pictorial 24h recall compared to structured observation for hygiene and sanitation behaviours (n pairs = 88)

| Behaviour | Observed agreement | Kappa -score | McNemar's test *p*-value | Reported only (n) | Observed only (n) | Reported and observed (n) | Sensitivity | Specificity | PPV | NPV |
| --- | --- | --- | --- | --- | --- | --- | --- | --- | --- | --- |
| Any handwashing with soap practiced | 70.5% | -0.054 | 0.009 | 20 | 6 | 1 | 14% | 75% | 4.8% | 91% |
| Any opportunities for child faeces disposal | 78.4% | -0.002 | 0.069 | 14 | 5 | 1 | 17% | 83% | 6.7% | 93% |
| Any safe child faeces disposal practiced | 80.7% | 0.024 | 0.049 | 13 | 4 | 1 | 20% | 84% | 7.1% | 95% |

Using structured observation as the reference group, sensitivity of P24hR was low for HWWS (14%), while specificity was much higher (75%). This resulted in a very low PPV of P24hR compared to direct observation but high NPV (Table 3).

### Measurement of child faeces disposal opportunities and behaviours

P24hR detected 16 total opportunities for child faeces disposal compared to 6 in structured observations, and safe disposal was recorded at all but one of these opportunities for each method (Table 2). Similarly to HWWS, the kappa statistics of the presence of any opportunities for child faeces disposal and presence of any safe child faeces disposal were both close to zero, indicating agreement no better than by chance, and a similar pattern of sensitivity and specificity resulted in a very low PPV (6.7% and 7.1%) and high NPV (93% and 95%) (Table 3).

## Discussion

This study evaluated the agreement between P24hR and structured observations and provides estimates of the reliability and validity of pictorial 24h recall as a novel method to measure hygiene and sanitation behaviours. Our findings suggest that P24hR has low agreement with direct observation, resulting in under-reporting of high frequency events, such as opportunities for handwashing, and over-reporting of “proper” or socially desirable behaviours, such as HWWS and safe child faeces disposal. Over-reporting by a self-reporting tool like P24hR is consistent with results from previous studies in Malawi (Chidziwisano et al., 2020) and other parts of the world (Curtis et al., 1993; Manun’Ebo et al., 1997). We found that P24hR tended to over-report handwashing opportunities when the average number of opportunities measured between methods was low, and under-report opportunities when the average number was high. This biphasic relationship illustrates that P24hR is a blunt instrument. The 95% LOA calculated (-7.62 to 4.89) are unacceptably wide considering the median 5 opportunities per participant observed. The high NPV suggests that P24hR is better suited for assessing the absence of specific behaviours rather than their presence, although the conceptual and practical utility of this may be limited.

Pictorial recall has been used in various other fields of research. In the field of nutrition, images representing different food groups and portion sizes have been widely used to facilitate dietary recall (Bulungu et al., 2021). Various tools have been validated to measure dietary diversity (Bulungu et al., 2021) and intake (Bulungu et al., 2021; Lazarte et al., 2012). Pictorial-assisted recall has been found to have high agreement with other methods of measuring time use in in low resource settings (Masuda et al., 2014). In water, sanitation, and hygiene research, pictorial aids have been used as a way to facilitate recall data, for example to measure water use (Esrey et al., 1992; Wright et al., 2006) or in daily diaries to measure diarrhoea episodes (Rego et al., 2021; Wright et al., 2006), but their measurement properties have not been fully evaluated.

Our study aligns with previous research that demonstrated that alternative methods for collecting self-reported hand hygiene behaviours are also subject to over-reporting. After adjusting for confounders, Schmidt and colleagues found that pictorial assisted estimates of HWWS were 13 percentage points higher than measuring through direct observation in a similar study population and 24 percentage points higher for post-defecation HWWS (Schmidt et al., 2019). However, differences between pictorial assisted recall compared to observation was smaller than the difference between traditional self-report and observations. In Ethiopia, Cotzen et al. (Contzen et al., 2015) compared covert script-based methods, in which respondents describe the sequence of actions between two events, to direct observation and traditional self-report methods to direct observation. While covert-script based methods had a higher correlation with observed behaviours than traditional self-report, they still over-estimated behaviours by 16 – 22 percentage points. Our study was not intended to compare P24hR against traditional self-reported behaviour; however, P24hR’s poor performance against structured observation by a variety of measures in this study makes any potential improvement against self-report of limited utility.

The strength of this study is that observations and P24hR were conducted on the same individuals only 24 hours apart, enabling direct comparison of two methods for measuring behaviour over the same approximate time period. A limitation of this study was the difficulty in accurately identifying the 6-hour observation period in 24h recall data. Despite collecting additional information to facilitate matching, some cut-off points had to be decided subjectively which may have resulted in misclassification of reported behaviours occurring before or after the time periods covered in the structured observations. The use of independent raters could be beneficial when isolating observation periods in recall data as well as measuring outcomes. Second, the schedule of data collection required P24hR to take place the day after observations, which lead to twelve participants being lost to follow-up. Given the poor performance of P24hR compared to structured observations in our analysis, it is unlikely that these 12 observations would have significantly improved the performance of P24HR. Third, child faeces disposal was rarely observed, which meant assessments were done using very few data points. Additionally, the high prevalence of null values for child faeces disposal and HWWS prevented the analysis of results through the Bland-Altman method. While the transformation of outcomes into categorical variables still permitted a relevant analysis of the data (Green, 2021), future tool evaluations could use negative binomial regression instead, as used by Schmidt et al., to prevent this issue (Schmidt et al., 2019). Finally, this study used structured observations as a reference. Observations have certain limitations, especially reactivity which could lead participants to wash their hands more than usual in the presence of an observer. However, observers are capable of precisely recording series of events and the timeframe in which they occur unlike P24hR. This means that opportunities and behaviours can be measured with less uncertainty.

### Conclusions

This study assessed the potential of pictorial 24h recall as a novel method to measure hygiene and sanitation behaviour for future evaluations of WASH interventions. Overall, agreement with structured observation was poor: P24hR tended to under-report hygiene opportunities and over-report socially desirable, “correct” behavioural outcomes. The negative predictive value of P24hR was high, although the conceptual and practical utility of this may be limited. While structured observations remain both time and resource intensive and may still result in biases, they remain the best method for measuring hygiene behaviours.

## Supporting information

P24hR Appendix Final

## Data Availability

All data produced in the present study are available upon reasonable request to the authors

## ^1^Abbreviations

P24hR: pictorial 24-hour recall
HWWS: handwashing with soap
TA: traditional authority
NGO: non-governmental organisation
LOA: limits of agreement
PPV: positive predictive value
NPV: negative predictive value

## Acknowledgements

The authors would like to thank the participants of our study for giving generously of their time. We also thank the field data teams who participated in both tool development and final data collection.

## Declarations

### Author contributions

Conceptualization: KC, RD; Design: OR, SB, BW, KC, RD; Acquisition of data: OR, BW, Analysis and Interpretation: OR, SB, RD; First draft of the manuscript: OR; Review and editing: SB, BW, KC, RD

