## Supplementary material for "Assessing the reliability and validity of pictorial-assisted 24-hour recall for measuring hand hygiene and child faeces disposal: a cross-sectional study in Malawi": P24hR Appendix Final

**Appendices**

**Appendix A: Additional information on diaries and photo cards.**

The P24hR tool was composed of two main elements: a series of pictures of daily activities and a diary sheet (Figure A1). The 24-hour diary sheet was divided up using temporal anchoring points rather than specific hours, as the study population rarely has access to clocks (Table A1). The list of daily activities was developed based on the list used by Schmidt et al. [9], adapted to the local context and pilot tested in two villages. After pilot testing, pictorial cards reflecting 41 distinct activities (Table A2) were identified. To facilitate their use, coloured symbols were added to each card to group into specific categories (for example, personal care activities all contained blue symbols while green represented activities around childcare) (Figure A1). The names, descriptions and drawings of activities can be found in Appendix B.

Figure A1. Finalized pictorial 24h recall tool

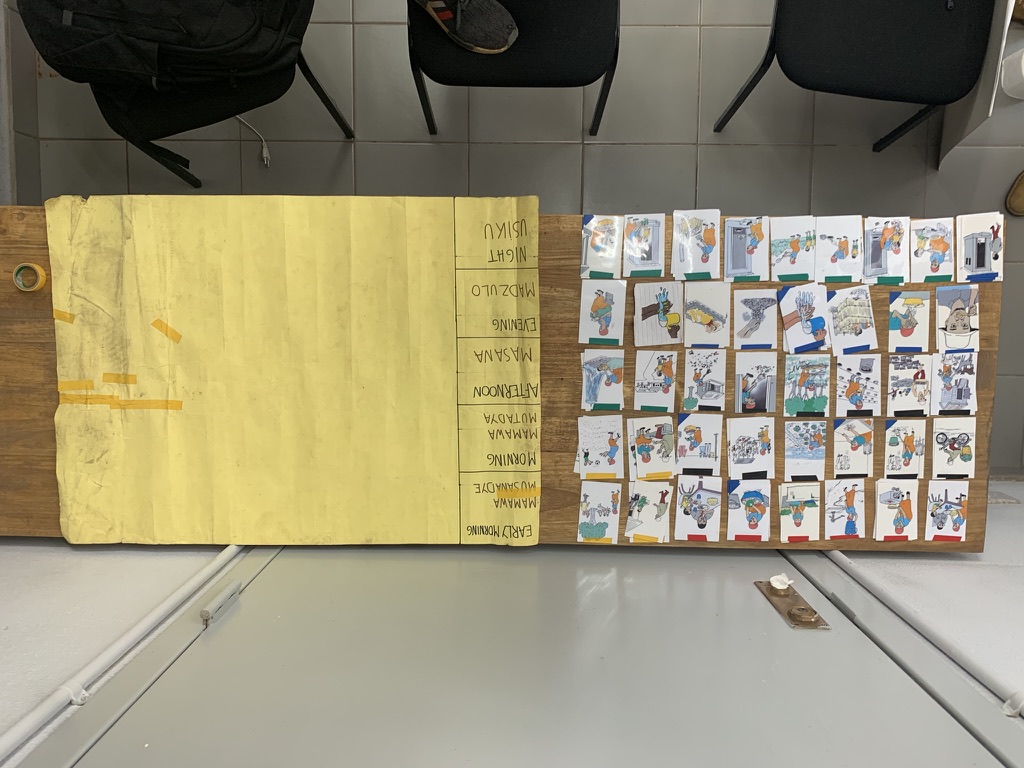

This picture includes the color-coded pictures of activities and the diary sheet on an A1 size piece of paper with time periods written in English and Chichewa.

Table A1: Temporal anchor points for diary sheets

| **Time period** | **Corresponding temporal anchor points** |
| --- | --- |
| Early morning | From the time the participant woke up until breakfast |
| Morning | From breakfast and through lunch |
| Afternoon | After lunch until sunset |
| Evening | After sunset until the evening meal |
| Night | After dinner until bedtime |

Table A2. List of activities by category

| Category | List of activities |
| --- | --- |
| Personal care | Going to the toilet  Wash face and body  Handwashing with water only*  Handwashing with water and soap*  Handwashing ash/mud/soil*  Brushing teeth  Sleep/rest |
| Work | Work in field  Tend to garden  Sell goods  Clean house  Clean house with water  Clean house surfaces  Clean house surroundings  Tend to animals  Casual work  Buy products  Wash clothes  Collect fire materials |
| Childcare | Give child bath  Take child for defecation  Clean child after defecation  Dispose/rinse of child faeces in latrines*  Dispose of child faeces in open*  Burry child faeces*  Throw child faeces into garbage*  Take child to/from school  Feeding child  Breastfeeding  Put child to bed |
| Food/ water | Eat (breakfast, lunch, dinner)  Wash food  Prepare food/drink  Serve food  Clean dishes  Collect water |
| Leisure | Visiting friends/relatives  Physical activity  Socialize  Pray |
| Other | For relevant activities completed by participants which were not specified in the list. |

*Target hygiene and sanitation behaviours.

### Appendix B. Description of activities with their pictures

| **Activity** | **Description** |
| --- | --- |
| **Personal care** | |
| 1. Going to the toilet | Respondent went to the toilet to defecate and/or urinate. Applicable even if respondent didn’t use a latrine.  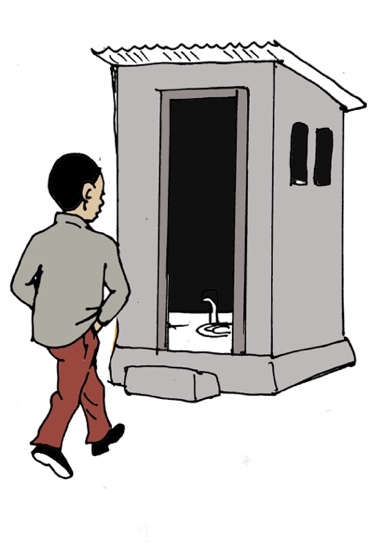 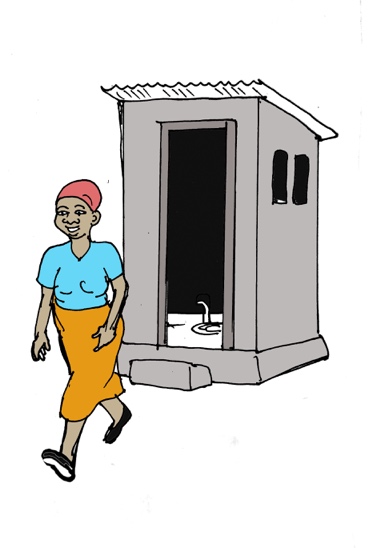 |
| 2. Wash face and body | Respondent washed their face and/or body with water (and soap if applicable)  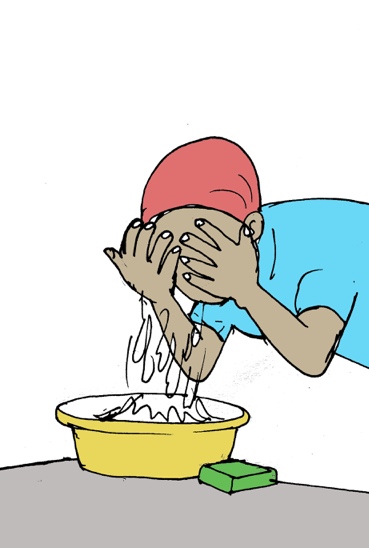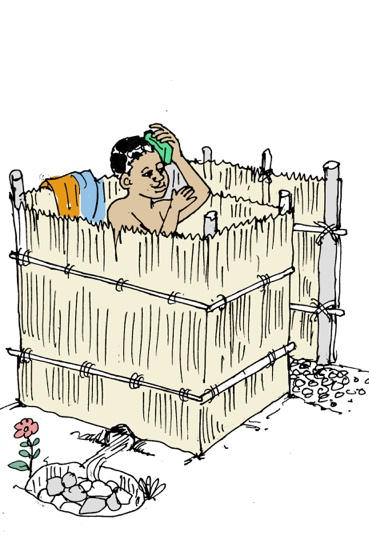 |
| 3. Handwashing with water only | Respondent washed their hands with only water  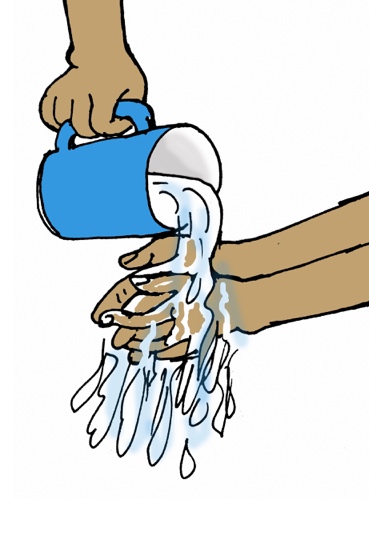 |
| 4. Handwashing with water and soap | Respondent washed their hands with water and soap  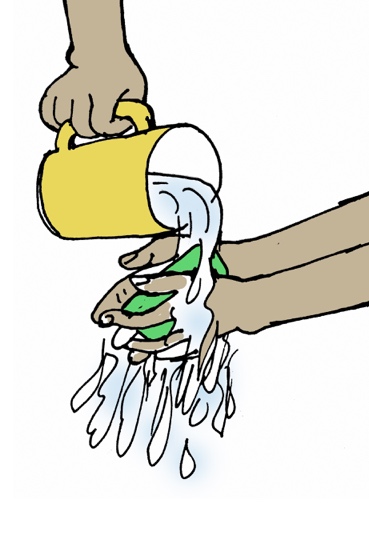 |
| 5. Handwashing ash/mud/soil | Respondent washed their hands with other material such as ash, mud or soil  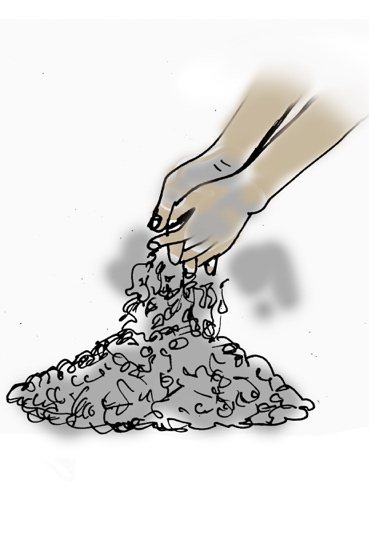 |
| 6. Brushing teeth | Respondent brushed their teeth with or without toothpaste  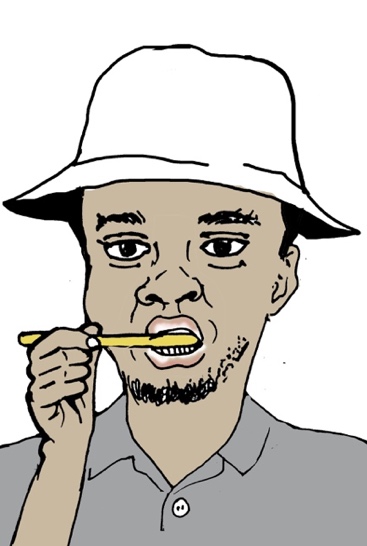 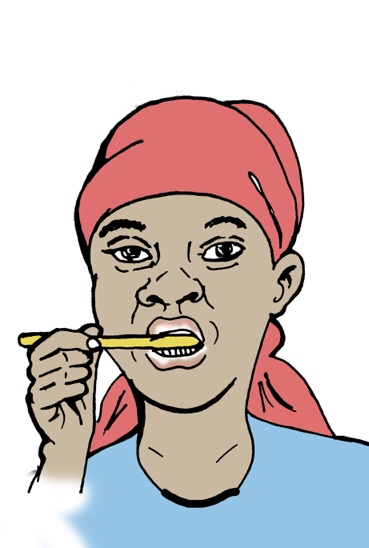 |
| 7. Sleep/rest | Respondent slept at night (end of the day)  Or respondent rested during the day either by laying down, relaxing, or taking a nap.  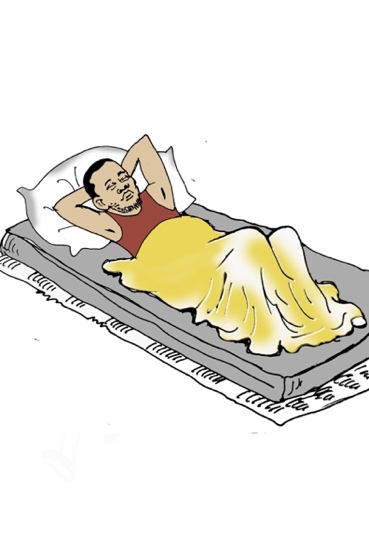 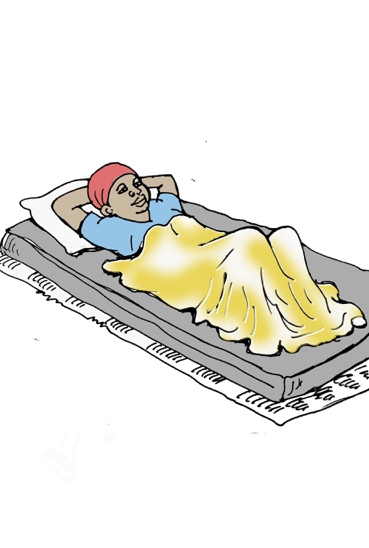 |
| **Work** | |
| 8. Work in field | Respondent worked in a field in which crops are not irrigated and depend on rainwater. Activities could be sowing, ploughing etc.  This includes working on own field or field of others.  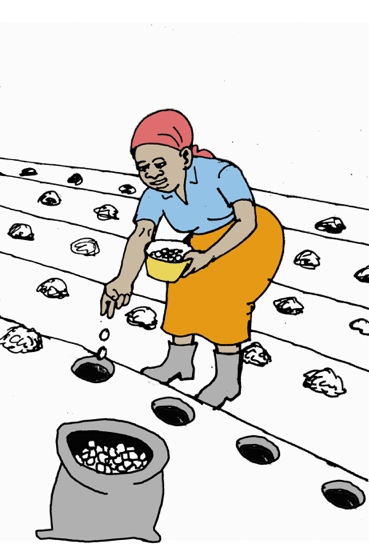 |
| 9. Tend to garden | Respondent tended to own garden in which vegetables are irrigated.  Activities include irrigation, picking vegetables etc.  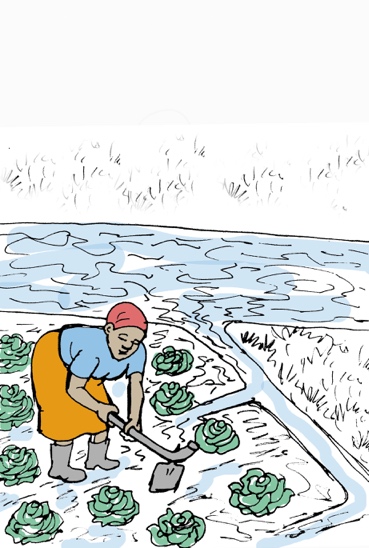 |
| 10. Sell goods | Respondent left the house to go sell products in places such as a market, village centre or side of road.  Goods being sold can be own production (surplus of harvest or vegetable garden) or can be products bought from providers are re-sold.  Businesses of any size are included in this category.  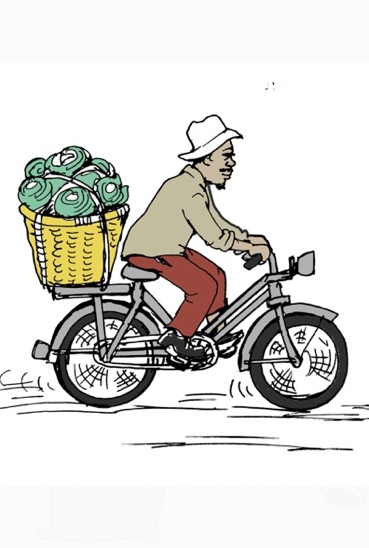 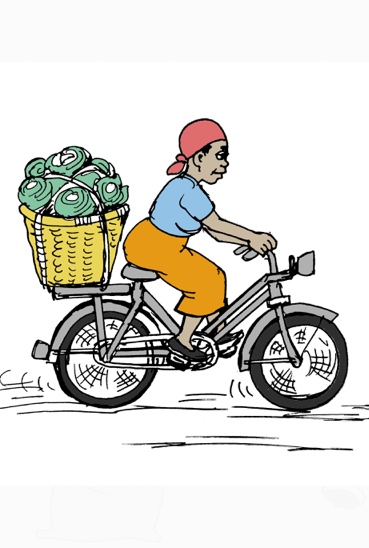 |
| 11. Clean house | Respondent cleaned (e.g. sweeping) the inside of their house including the porch (area connected to the house and protected by the same roof as the house).  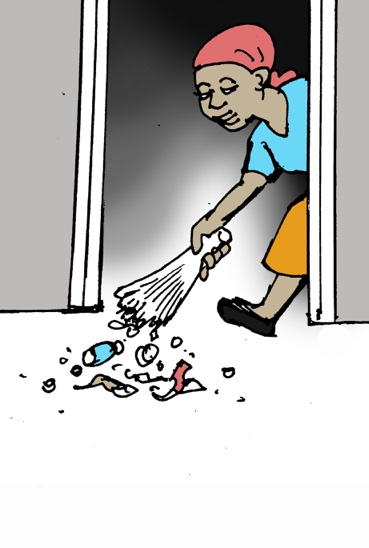 |
| 12. Clean house with water | Respondent cleaned the inside of their house 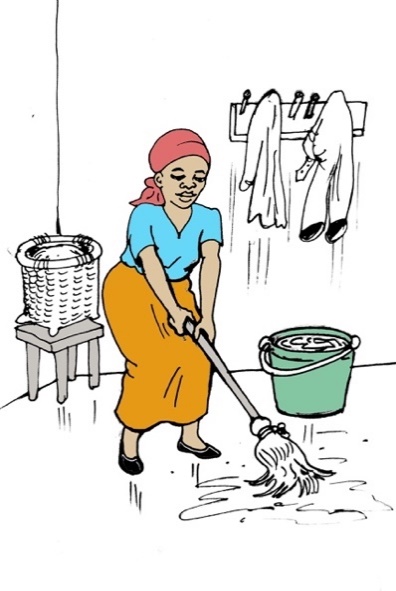with water (e.g. mopping) |
| 13. Clean house surfaces | Respondent cleaned surfaces inside their homes (e.g. tables or furniture)  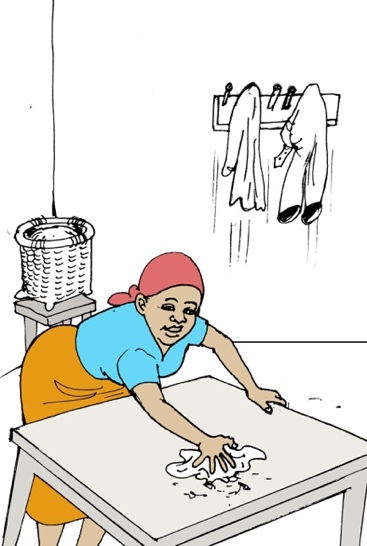 |
| 14. Clean house surroundings | Respondent cleaned (e.g. sweeping) the outside of the house, meaning the immediate surroundings.  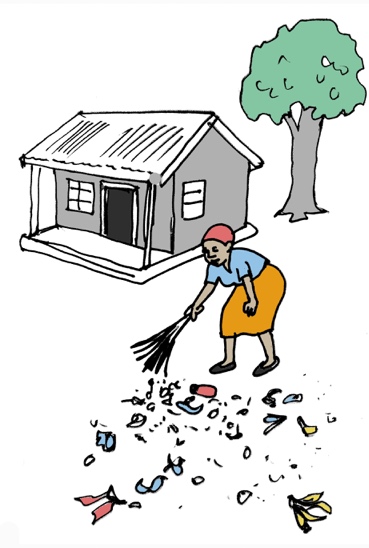 |
| 15. Tend to animals | Respondent either fed, fetched some animal feed in nature (not bought it somewhere), gave animal water, took out animal to graze, cleaned animal and/or animal house.  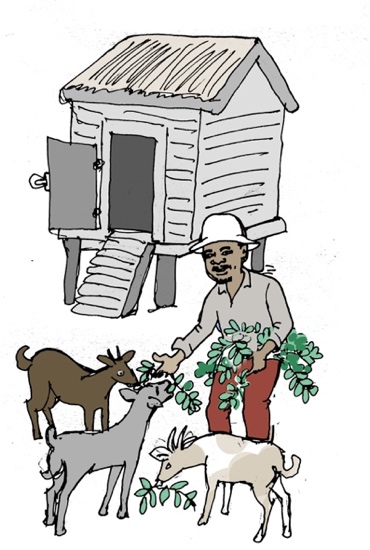 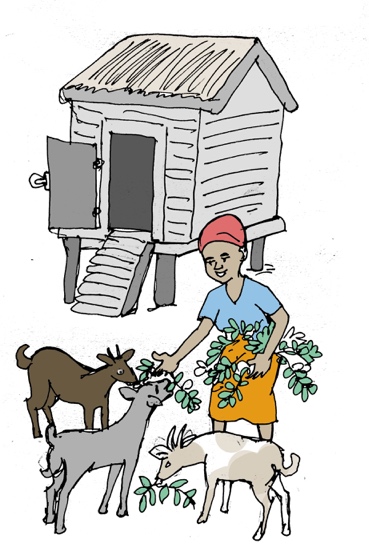 |
| 16. Casual work | Respondent worked for others on various tasks such as collecting water to make bricks.  This excludes field work, even if person is doing field work for someone else.  This can include more formal works as well.  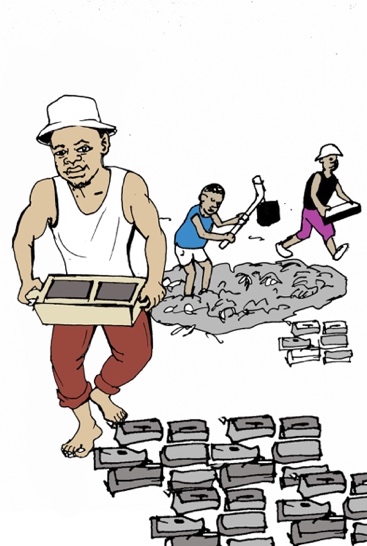 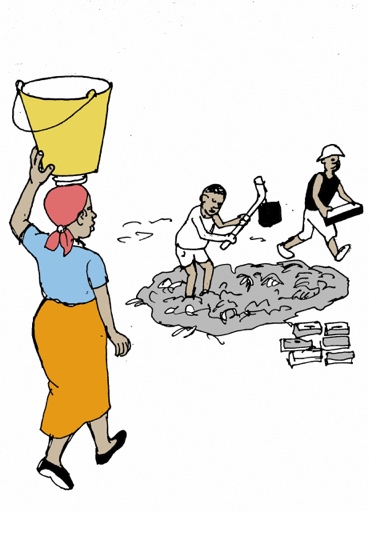 |
| 17. Buy products | Respondent left the house to buy foodstuff (at a market or elsewhere) as well as other products such as clothes, electronics etc (at a market or elsewhere)  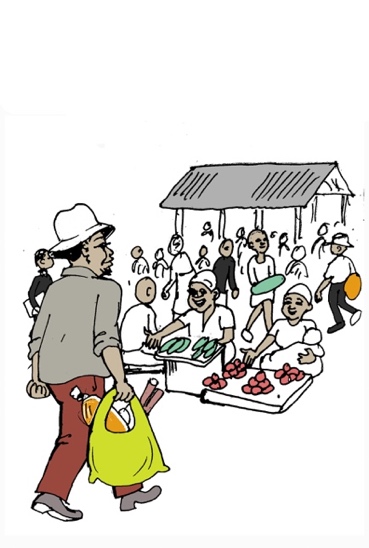 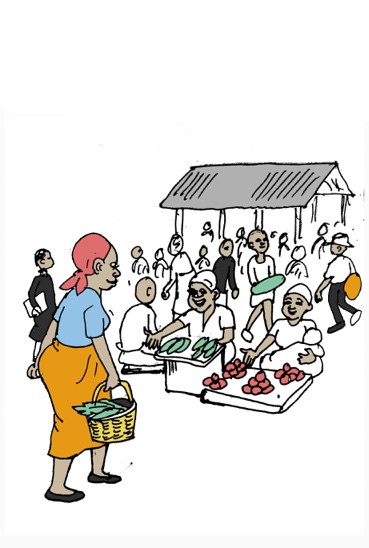 |
| 18. Wash clothes | Respondent washed and dried clothes including storing them once dried.  This includes their own clothes or other household members’ clothes such as children.  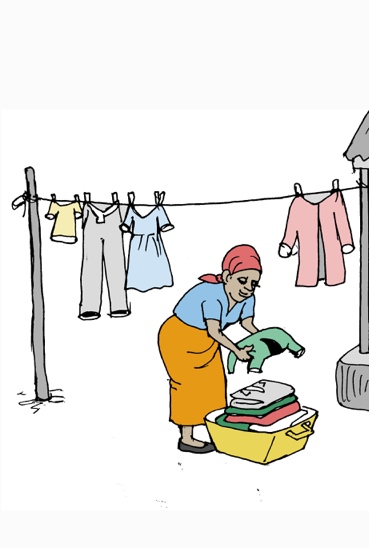 |
| 19. Collect fire materials | Respondent left the house to go collect firewood or briquettes.  This excludes buying firewood (see 15).  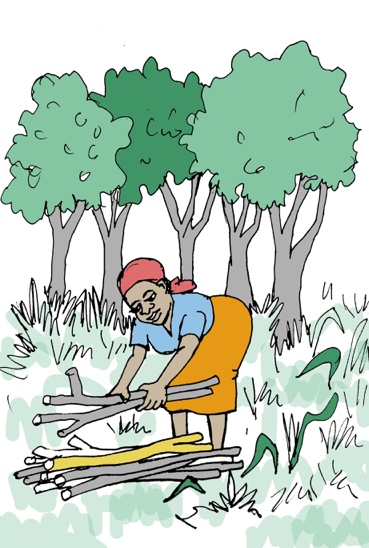 |
| **Childcare** | |
| 20. Give child bath | Respondent gave children (under 18 of age) a bath with or without water.  Excludes children bathing themselves alone and cleaning child’s bottom because they defecated (see 20).  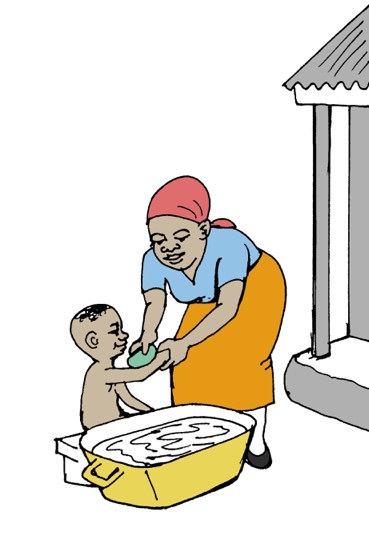 |
| 21. Take child for defecation | Respondent accompanied child to place of defecation (toilet, field, bush) and help him/her getting dressed/undressed to defecate.  This excludes child defecating in diaper.  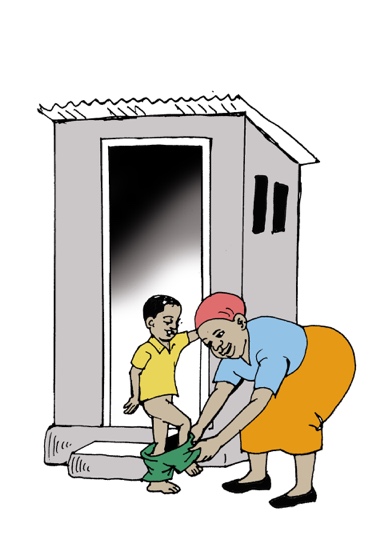 |
| 22. Clean child after defecation | Respondent cleaned child after defecation with water and/or changed diapers.  This includes washing the diaper so that child has a clean one.  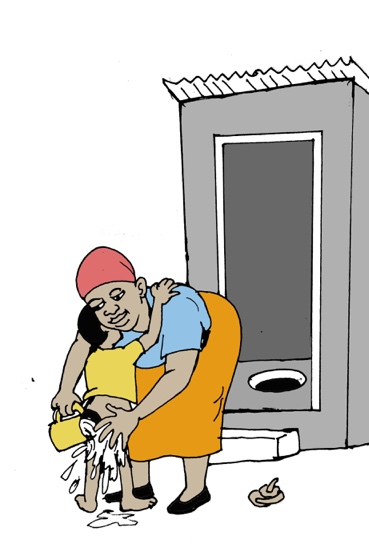 |
| 23. Dispose/rinse of child faeces in latrines | Respondent disposed (threw away) child faeces in toilets including rinsing diaper in toilet or picking up faeces from place of defecation (other than toilet) and putting into toilet. This excludes throwing away diapers in toilets.  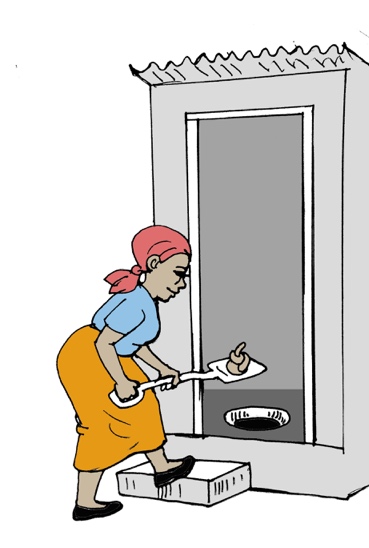 |
| 24. Dispose of child faeces in open | Respondent disposed of child faeces/ dirty diapers in bush, field, drain or ditch. This includes not disposing of them and disposing of diapers in latrines.  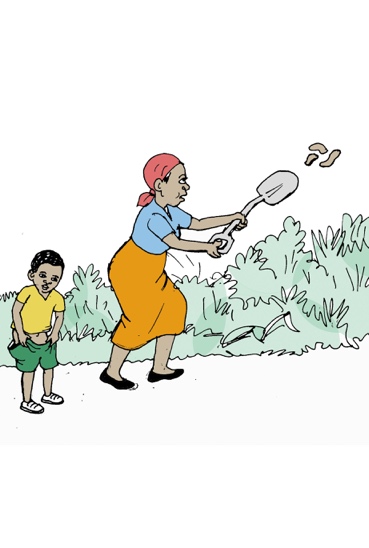 |
| 25. Burry child faeces | Respondent disposed of child faeces by burying them.  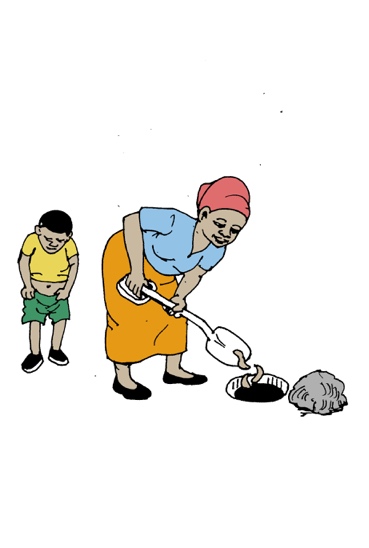 |
| 26. Throw child faeces into garbage | Respondent threw child faeces/dirty diapers away in same bin as the rest of solid waste.  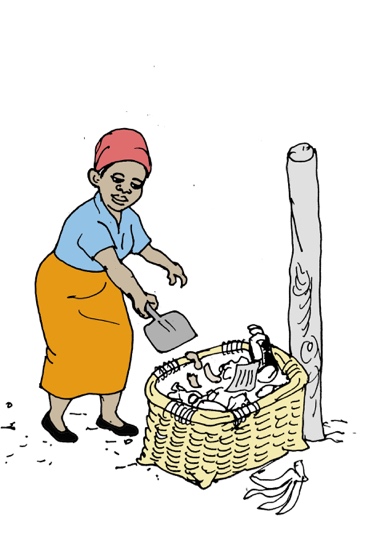 |
| 27. Take child to/from school | Respondent accompanied child to school and picked up child from school.  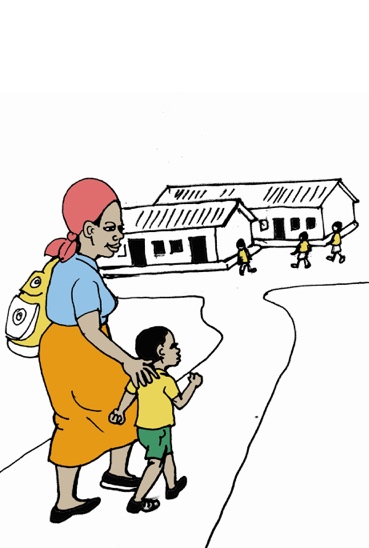 |
| 28. Feeding child | Respondent fed child at a different moment than the one at which they ate.  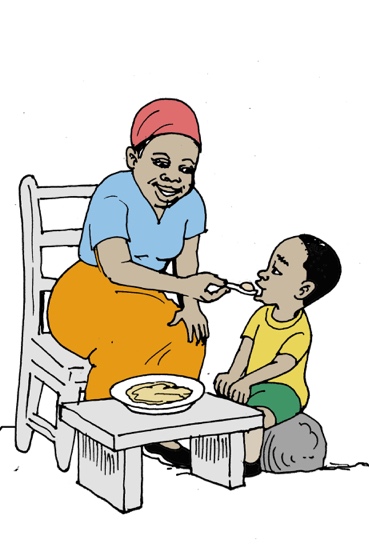 |
| 29. Breastfeeding | Respondent breastfed child.  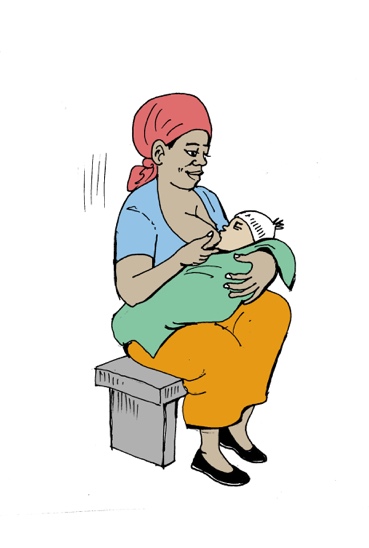 |
| 30. Put child to bed | Respondent prepared the bedroom (prepare mattress, mosquito net) and put child to bed at different time than the one at which they went to bed. This includes putting child down for a nap.  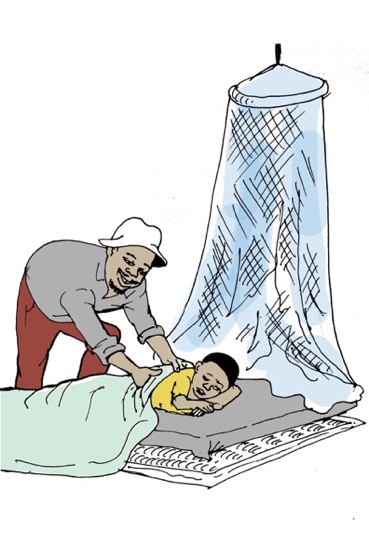 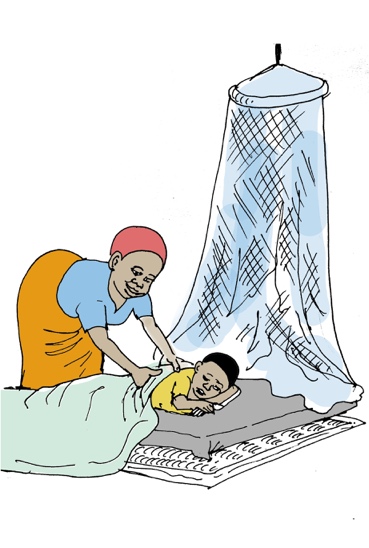 |
| **Food/drink** | |
| 31. Eat (breakfast, lunch, dinner) | Respondent ate breakfast, lunch, or dinner.  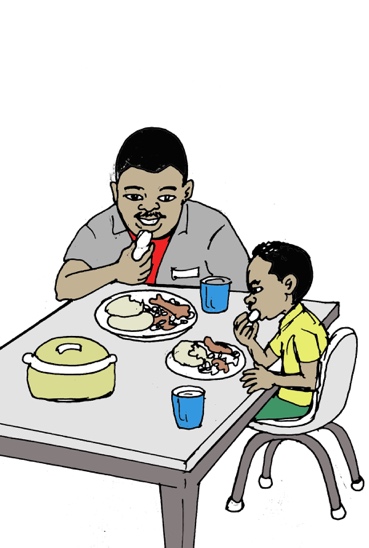 |
| 32. Wash food | 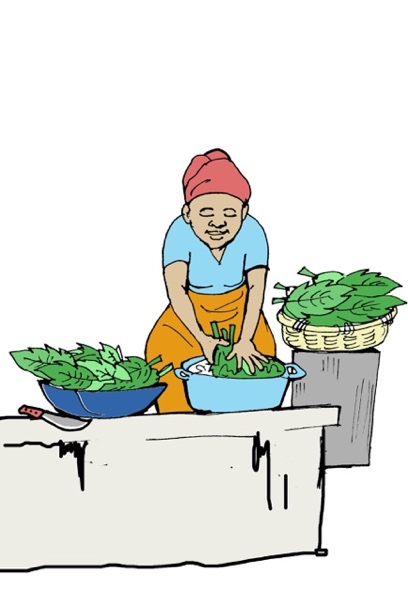Respondent washed their food (vegatiables, fruits, …). |
| 33. Prepare food/drink | Respondent prepared food for themselves and/or others. Same as cooking. This includes heating water for consumption. This excludes preparing food only for children (see 27).  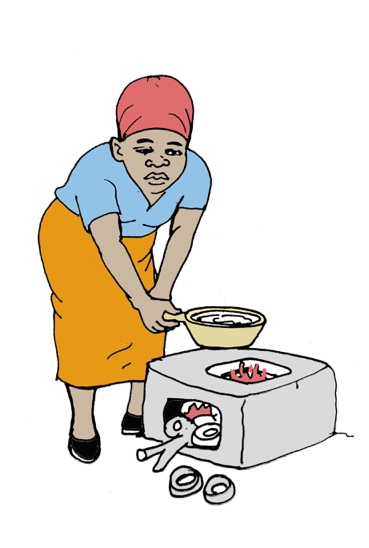 |
| 34. Serve food | Respondent served food to other people but didn’t eat themselves.  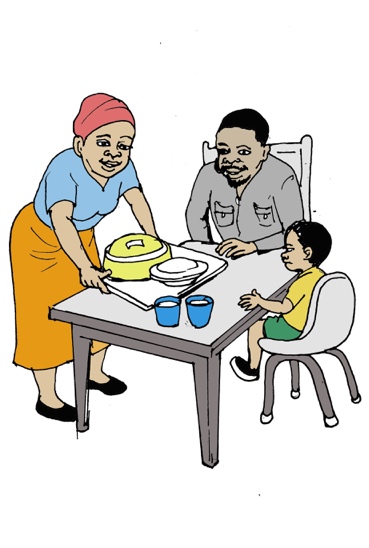 |
| 35. Clean dishes | Respondent cleaned utensils, cups, plates, cutlery (i.e. anything used to eat or drink)  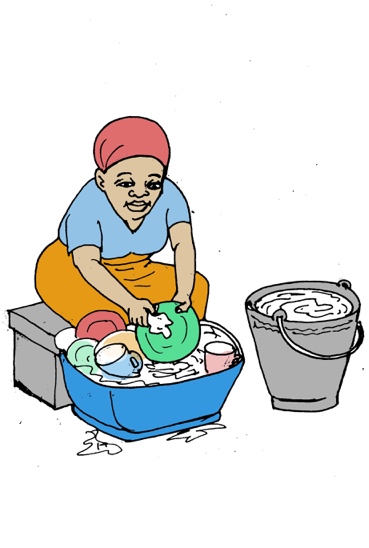 |
| 36. Collect water | Respondent left the house premises to go collect water for drinking/cooking, washing (face, body, hands) or cleaning (utensils, house)  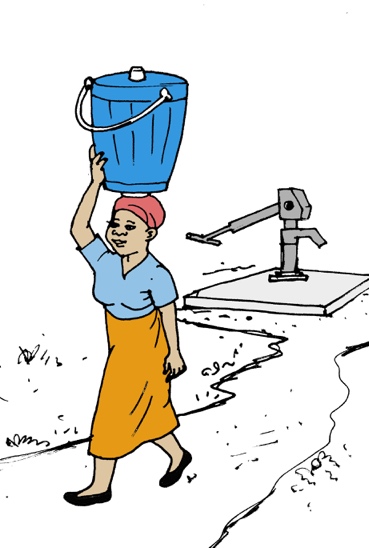 |
| **Leisure** | |
| 37. Group gatherings (“going out”) | Respondent left the house to go meet other people or participate in a social activity such as church, drinking joints, movies, watch football, hangout with friends.  These activities should all take place in public spaces (not someone’s house).  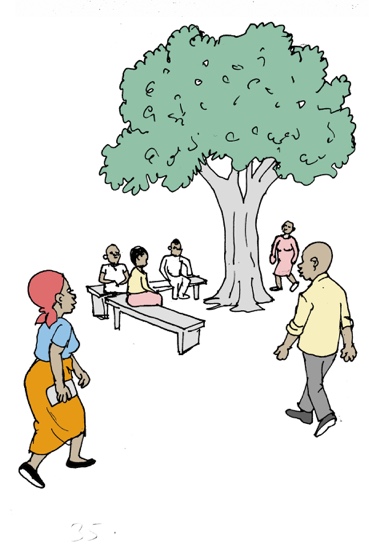 |
| 38. Visiting friends/relatives | Respondent left the house to go visit friends/relatives in a private space (friend/relative’s house for example)  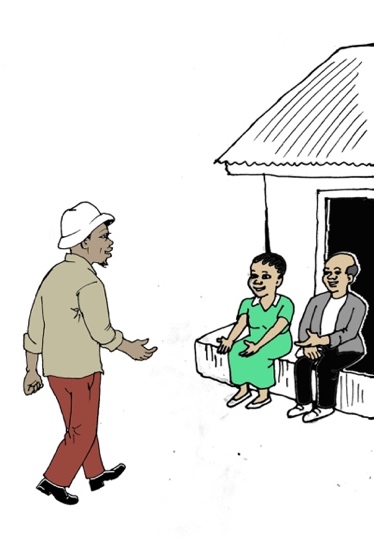 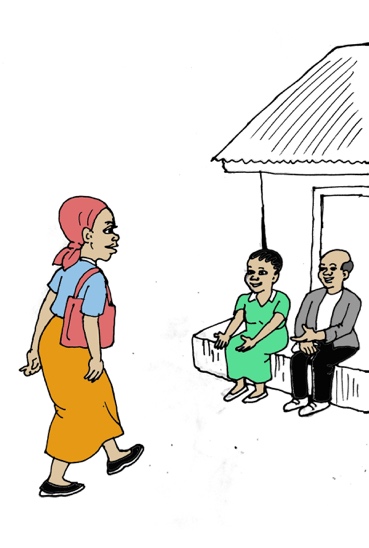 |
| 39. Physical activity | Respondent left the house to go do a physical activity such as play sports (eg: play football) or go out for a walk (for leisure)  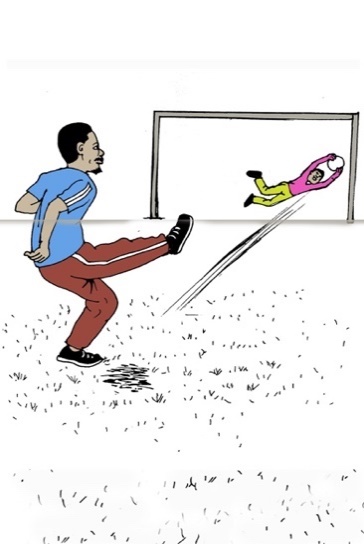 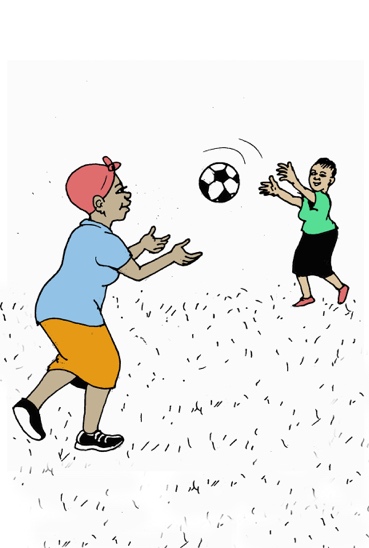 |
| 40. Socialize | Respondent was visited by people or played games with people (cards/bawo) while remaining at their house.  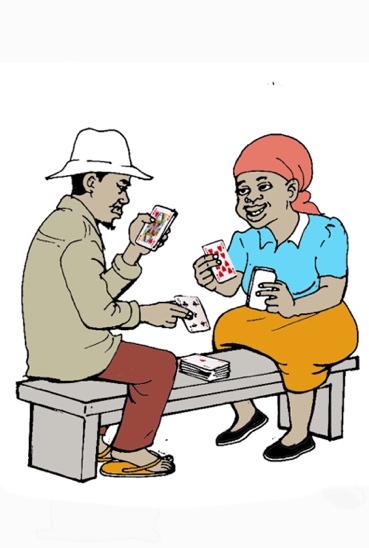 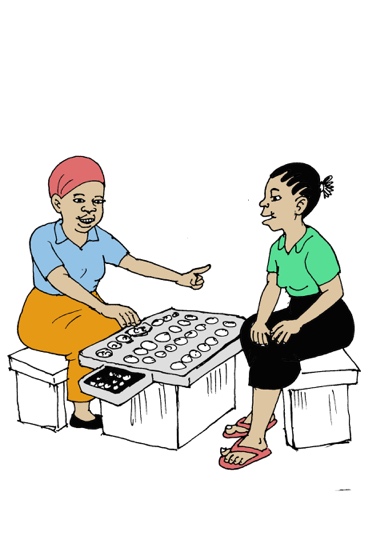 |
| 41. Other | Any activity that respondent did that is not included in the ones above. Only use as last resort. Specify what activity it was. |
